# Risk factors for renal stone development in adults with primary hyperparathyroidism: A protocol for a systematic review and meta-analysis

**DOI:** 10.1101/2025.01.22.25320716

**Authors:** Mohammad Jay, Sorina Andrei, Peter Hoang, Hussein Samhat, Roland Jones, Rui Fu, Lorraine Lipscombe, Antoine Eskander

## Abstract

**Background:** Primary hyperparathyroidism (PHPT) is characterized by overactive parathyroid glands. Renal stones (RS) are a common complication of PHPT and is associated with increased morbidity. However, the risk factors for RS in PHPT are not well-established and the latest international PHPT guideline highlights the need for further research into this area.

**Objective:** We aim to summarize and meta-analyze the existing evidence on prespecified risk factors associated with RS in adults with PHPT.

**Methods and Analysis:** We will search MEDLINE, EMBASE, and Cochrane Central from inception. Two independent reviewers will screen studies and include prospective/retrospective cohort, case-control, and cross-sectional designs in adults (≥18 years) with PHPT. Randomized trials, conference abstracts, case reports, and commentaries will be excluded. Two reviewers will independently extract data on population characteristics, risk factors, RS outcomes, and assess risk of bias using the Quality in Prognostic Studies tool. A random-effects model will be used to pool odds ratios. We will separately pool adjusted (primary analyses) and unadjusted odds ratios (secondary analyses) with their corresponding 95% confidence intervals. Certainty will be evaluated with the Grading of Recommendations Assessment, Development, and Evaluation framework. Heterogeneity will be assessed using the I² statistic and publication bias will be evaluated with funnel plots.

**Discussion:** Early identification of patients with PHPT at high risk for RS can facilitate the implementation of preventive strategies and reduce morbidity. Furthermore, recognizing these risk factors can assist clinicians in prioritizing treatment for those at higher risk, ultimately improving patient outcomes.

**Protocol registration:** The protocol was registered in PROSPERO on November 14, 2024 (registration ID: CRD42024608180).

**Funding:** No source of financial funding was used.

## 1. Introduction

Primary hyperparathyroidism (PHPT) is characterized by elevated serum parathyroid hormone (PTH) levels and consequent disruption of calcium metabolism (1, 2). The resulting hypercalcemia can lead to several serious complications, including renal stones (RS), renal failure, and osteoporosis (3). PHPT affects roughly 600,000 Canadians (2, 3). An estimated 40% of patients with PHPT develop RS, a complication associated with severe pain leading to frequent emergency department (ED) visits, a 1.4-fold increased mortality risk, and a 2.3-fold higher risk of renal failure (4, 5). In many countries, the direct costs of RS management and lost work productivity is estimated to total billions of dollars annually (6, 7).

PHPT treatment guidelines recommend reserving curative treatment, parathyroidectomy, for patients at high risk of complications, such as RS (8–10). Early identification of patients with PHPT at high risk of developing RS has important implications for optimizing monitoring schedules, implementing preventative strategies and prioritizing parathyroidectomy (11–15). Although multiple systematic reviews have examined RS risk factors in the general population, their applicability to PHPT is uncertain, given that this population experiences disease-specific changes in calcium metabolism (16–19). For instance, older age, male sex, and hypercalciuria are commonly recognized risk factors in broader RS literature, but whether these factors have similar associations in individuals with PHPT is unclear (18). Equally important is whether PHPT-specific drivers of incident or recurrent RS—such as excessively high serum calcium or PTH— operate independently or in conjunction with known risk factors from the general population (16, 20, 21). Most evidence to date has focused on the hallmark biochemical features of PHPT (i.e., serum calcium and PTH levels) and patient demographic characteristics such as age and sex (16, 20, 21). However, findings across studies have been inconsistent (21); while some studies suggested that RS risk is associated with younger age, higher serum calcium and higher urine calcium, other studies did not replicated these findings (16, 22–27). Studies which found different RS risk factors in PHPT compared to the general population suggest that RS risk profile in PHPT is unique (21, 24, 27).

Further complicating our understanding, emerging data suggest that serum vitamin D levels may also play a role in RS risk among patients with PHPT (28, 29). Vitamin D deficiency is common in patients with PHPT and maintaining normal vitamin D levels are essential for bone health (30). Vitamin D has been associated with higher risk of RS.(29) Conversely, as vitamin D intake increases calcium absorption, it may exacerbate hypercalcemia and hypercalciuria in some patients, potentially increasing RS risk (28).

Recognizing these knowledge gaps, the latest international PHPT guideline highlights “[renal] stone risk” as a research priority, underscoring the need to establish evidence-based risk stratification in this population (13). We therefore aim conduct a systematic review and meta-analysis to consolidate and critically appraise the available evidence on the association of six *a priori* defined risk factors—age, sex, urine calcium, serum calcium, serum PTH and serum vitamin D—with both incident and recurrent RS in PHPT. Although a narrative review in 2011 summarized the RS risk factors in PHPT, no systematic review or meta-analysis has been conducted on this topic (21). By integrating data from a broad range of study designs and settings, we aim to provide a clearer understanding of which patients with PHPT are most susceptible to RS development, ultimately informing clinical decision-making.

## 2. Objectives

The primary objective of our review is to summarize and meta-analyze the existing evidence on the association of five risk factors (sex, age, serum calcium, urine calcium, and PTH) with both incident (new) or recurrent RS development among adults (≥18 years old) with PHPT in outpatient or inpatient settings up to 10 years before or after PHPT diagnosis. For each risk factor, the comparator will be an alternative category of the risk factor. The secondary objective will include evaluating the association of these six risk factors with recurrent RS development.

## METHODS

### 3. Study Design

Our systematic review will be conducted in accordance with the guidelines outlined in the Cochrane Handbook for Systematic Reviews of Interventions (31). This protocol was developed following the Preferred Reporting Items for Systematic Reviews and Meta-Analyses (PRISMA) 2015 checklist (S1. Appendix) (32) and was registered with PROSPERO: an international prospective register of systematic reviews on November 14, 2024 (registration ID: CRD42024608180) (33). Data screening has been completed. Data extraction is estimated to be completed by March-April 2025 and complete results are expected by April-May 2025. The final systematic review will follow the PRISMA-2020 reporting guidelines (34).

## 4. Eligibility Criteria and Outcomes

### Study Type

We will include full-length peer-reviewed prospective cohort, retrospective cohort, case-control, and cross-sectional studies. For cohort studies, the starting point (time 0) will be the diagnosis of PHPT, with follow-up measuring the development of RS. We will not restrict the time duration between PHPT diagnosis and RS development as our primary outcome is RS occurrence irrespective of the time of PHPT diagnosis. Additionally, we will not apply any language, geographic or time restrictions to the search. Google Translate will be used to translate non-English studies if necessary (35). Studies will have to report sufficient data to allow for directly or indirectly estimating odds ratios (OR), relative risks (RR), or hazard ratios (HR) with 95% confidence intervals (CIs) for at least one of the predefined risk factors.

We will exclude studies that only include paediatric patients (<18 years old), case series (i.e., those with no quantitative data or fewer than five patients), case reports, commentaries, editorials, non-human studies, review articles, and conference abstracts. Clinical trials will also be excluded to avoid the potential for random allocation obscuring the association between the risk factors and RS (36).

### Participants

Adults (≥18 years old) with symptomatic or asymptotic PHPT will be included. Studies presenting data for both adults and children will only be included if the relevant data are presented separately for adults. The diagnostic criteria for PHPT will be based on internationally recognized standards (13, 37). As most guidelines exclude normocalcemic PHPT in the formal definition of PHPT, we will exclude these patients (13, 37). Studies presenting data on both hypercalcemic and normocalcemic PHPT patients will only be included if the data for these groups are reported separately. We will also exclude data obtained after parathyroidectomy to avoid its confounding effect on RS risk. Studies that include patients who have undergone parathyroidectomy will only be considered if they report relevant data collected before the procedure (12).

### Index prognostic factor

Studies must include at least one of the following a-priori risk factors: 1-age, 2-sex, 3-urinary calcium, 4-serum calcium, 5-serum PTH, 6-serum vitamin D.

### Comparator

For each risk factor, the comparator will be patients in an alternative category of the risk factor.

### Outcome: 1)

**The primary outcome** will be incident and recurrent RS development during any time frame relative to PHPT diagnosis. This outcome will provide a comprehensive assessment of RS burden in PHPT, capturing both new and recurrent RS events to characterize the full spectrum of RS. The lack of established risk factors and systematic reviews in this area underscores the need for robust data on the overall risk and natural history of RS in PHPT (15, 38–40). 2) **The secondary outcomes** will be recurrent RS development. Given that prior RS are a strong predictor of recurrence, clinicians often adopt more intensive management for these patients, including closer monitoring and higher likelihood of parathyroidectomy (39, 41). Focusing on recurrent RS will clarify whether a “one-size-fits-all” approaches to RS risk factors are adequate or if tailored prevention strategies are required for these higher-risk patients. 3) As an **exploratory outcome**, we will assess the mean difference of continuous risk factors between RS and no-RS groups to provide a clinically meaningful interpretation (42, 43). The definition of RS will be based on internationally recognized standards, primarily based on the presence of symptoms or radiographic findings (7, 44, 45).

### Timing

We will assess RS occurrence within a 10-year window before or after PHPT diagnosis as reported in the included studies. This 10-year period is standard in RS risk studies, effectively balancing the capture of relevant clinical outcomes with the minimization of confounding factors (25, 46, 47).

### Setting

*Any* healthcare (e.g., inpatient, outpatient), geographic (e.g., urban, rural), and study-specific (e.g., single-centre, population based) will be included.

## 5. Information Sources

The following online databases will be searched from their inception: Medline (OVID interface, 1946 onwards), Embase (OVID interface, 1947 onwards), and Cochrane Central Register of Controlled Trials (Wiley interface, current issue). References of all eligible articles will be reviewed for additional studies meeting the eligibility criteria (backward citation searching) (48). Databases containing grey literature will not be searched as our focus is on peer-reviewed studies.

## 6. Search Strategy

Literature search strategies will be developed using a combination of subject headings (MeSH, EMTREE) and keywords. Database searches will be conducted with the aid of an experienced health information librarian. Two concepts will be included: 1-primary hyperparathyroidism, and 2-RS. These two concepts will then be combined using the “AND” operator to obtain final search results. We will limit the search to only studies involving human subjects and will not apply any language restrictions.

A preliminary search strategy is provided in S2. appendix.

## 7. Study Records

### 7.a. Data Management

All citations from the literature search will be imported into Covidence and duplicate studies will be removed (49). Following the completion of screening, a PRISMA flow diagram will be generated using Covidence (50).

### 7.b. Selection Process

Two reviewers will independently screen titles and abstracts of studies uploaded in Covidence, applying prespecified eligibility criteria. The full texts of studies deemed potentially relevant will then be independently reviewed for inclusion in the final analysis by the same reviewers. Reasons for excluding studies will be recorded. Reviewers will resolve disagreements by discussion, and a third reviewer will adjudicate the unresolved disagreements. A calibration exercise involving a small sample of studies (e.g., three) will be conducted before screening begins to ensure consistency and refine eligibility criteria. Reviewers will not be blinded to journal titles, study authors or institutions.

### 7.c. Data Extraction

For each included study, two reviewers will extract data independently and in duplicates and record the data in standardized data collection tables in Microsoft Excel. These tables will be organized in the recommended PICOTS format for prognostic studies (patient, index prognostic factor, comparator prognostic factor, outcome, time and setting) (51), and will initially be piloted on three studies to allow necessary adjustments for accuracy and completeness. The tables will be created using a modified version of the checklist for critical appraisal and data extraction for systematic reviews of prediction modelling studies for prognostic factors (CHARMS-PF)(52, 53). Reviewers will compare the collected data, resolve disagreements by discussion, and a third reviewer will adjudicate the unresolved disagreements.

## 8. Data Items

We will record the following data from the included studies where available: 1-Basic study details: first author’s last name, publication year, study design, country of origin, sample size, setting, data sources (e.g., self-report, chart review, administrative data), follow-up duration, and loss to follow-up, funding sources. 2-Patient Demographics: age and sex. 3-Biochemical Risk Factors: 24-hour urine calcium, total serum calcium, serum ionized calcium, serum PTH, 25-hydroxy vitamin D, and 1-25 dihydroxy vitamin D. Units of measurement for all biochemical variables and the time at which the patient demographics and the biochemical risk factors were assessed in relation to RS development will be recorded. 4-Clinical History: time of PHPT diagnosis in relation to RS development, and criteria for PHPT diagnosis. 5-Outcome Measures: Presence of RS (Specify if RS are incident or recurrent), criteria for RS diagnosis. 6-Other Relevant Clinical Information: osteoporosis and treatment with calcium supplement (S3. Appendix).

Whenever available, we will extract adjusted and unadjusted OR, RR, or HR, and their measures of variance. If these measures of association are not available, the relevant descriptive statistics (e.g., mean, median) will be recorded to estimate the unadjusted OR (54, 55). We will also record the variables used to adjust for each risk factor. We will collect data on all *a priori* defined risk factors and outcome-compatible information, regardless of RS definition or timing of RS diagnosis. We will convert data from different studies into consistent units of measurement to ensure uniformity.

### Missing Data

In cases of missing or unclear data, authors will be contacted up to two times, allowing two weeks for a response per attempt. If no responses are received regarding essential study eligibility criteria, the study will be excluded (56). Similarly, if essential data required for analysis are not obtained, the study will be excluded from the relevant analysis but will contribute to the descriptive analyses (57).

## 9. Quality Appraisal

Two reviewers will independently assess each risk factor’s risk of bias using the Quality in Prognostic Studies (QUIPS) tool. Reviewers will resolve disagreements through discussion, and a third reviewer will adjudicate the unresolved disagreements. This tool evaluates six domains: study participation, study attrition, prognostic factor measurement, outcome measurement, study confounding, and statistical analysis and reporting (58). Each domain will. be rated as high, moderate, or low risk of bias (58). This tool will be initially piloted using three randomly selected studies to confirm consistency across reviewers. A study will be deemed low risk overall if all domains are rated as low risk. Conversely, it will be considered high risk if at least one domain is rated high. Any ratings between these categories will be rated as moderate risk of bias (59, 60). Traffic-light plots will be generated using R-Version 4.4.1.

## 10. Data Synthesis

We will conduct meta-analyses on all prespecified risk factors and outcomes reported by at least two studies, using the generic inverse variance method within a random-effects model to account for between-study heterogeneity (61, 62). To ensure robust estimation of uncertainty in the overall effect size, the Hartung-Knapp-Sidik-Jonkman adjustment will be applied when at least four studies are included in the meta-analysis (63). When fewer than four studies are available, the Wald method will be used instead (64). The restricted maximum likelihood method will be used to estimate between-study variance (65). Between-study heterogeneity will be assessed by visually inspecting forest plots for overlapping point estimates and CIs, and formally evaluated using the chi-square test and I² statistic (66). Forest plots, summary effect measures with 95% confidence intervals (CIs), and prediction intervals will be generated using R Version 4.4.1.

### Primary Analysis

We will meta-analyze adjusted risk factors. We will report summary adjusted odds ratios (aORs) with 95% CIs for the association with RS. We will pool all adjusted risk factors regardless of the adjustment variables, without requiring a minimum set of adjustment factors (55). We have chosen this approach for This approach to pooling adjusted effect estimates irrespective of the specific covariates is chosen for two primary reasons: 1) The risk factors and confounders for renal stones in PHPT are not well-established(67). Consequently, we expect the included studies to vary widely in the variables they adjust for. Pooling all adjusted estimates allows us to capture at least the effect of partial confounding control (68, 69). 2) Given the lack of established risk factors, our priority is establishing the direction of risk factors with adequate precision. Maximizing the number of studies in the meta-analysis enhances the precision of effect estimates. Previous systematic reviews on prognostic factors have adopted similar approaches (21, 55, 70, 71).

### Secondary Analysis

We will examine the unadjusted risk factors. For these, we will estimate summary unadjusted odds ratios (uORs) with 95% CIs for the association with RS (72), using the following stepwise approach: 1) **Direct Use of Unadjusted ORs:** If unadjusted ORs are reported by the study, these will be directly used in the meta-analysis. 2) **Conversion of Relative Measures**: If studies report relative risk (RR) or hazard ratio (HR), HR will equated to RR, and these measures will be converted to OR.(55, 73) 3) **Calculating ORs from Regression Coefficients**: When regression coefficients are reported without confidence intervals (CI), ORs and their variances will be calculated using the regression coefficients and p-values from logistic or linear regression.(74) If regression coefficients and CIs are reported, ORs will be calculated using the formula: OR= ebeta, where e= 2.718 and β is the regression coefficient (74, 75). **4) Calculating ORs from Cox Regression and Kaplan-Meier Curves:** When direct reporting is absent, we will use the number of events and the p-values from reported analyses such as log-rank tests or Cox regressions (54, 76). If only Kaplan-Meier curves are reported with no available HR, we will reconstruct the raw time-to-event data from Kaplan-Meier curves and use this data to estimate the HR (77). **5) Deriving ORs from 2×2 Tables:** If no measures of association are described but categories of a risk factor in relation to RS are reported, a 2×2 table will be constructed to calculate the OR. **6) Tertile Approach for Continuous Variables:** If neither measures of association nor categorical data are available, continuous variables will be analyzed by assuming a normal distribution (55, 78). Using the mean and standard deviation (SD), we will create three groups (tertiles) in R Version 4.4.1 and calculate the OR for the highest third versus the lowest third of each prognostic factor (55, 70, 79, 80). When only median and interquartile range (IQR) are reported, these values will be converted to mean and SD (79). If means and SDs are reported per group, data will be combined using the approach suggested in the Cochrane Handbook (81). Studies will be excluded if measures of variance are unavailable (82).

To ensure the assumption of normality and avoid unstable tertile conversions, we will apply the following criteria. A study will be included in the tertile analysis if at least two of the following three criteria were met: 1) Minimum number of participants in each group >20 (83). 2) Coefficient of variation (CV) between 0.12 and 0.6 for both groups (84–87). 3) Maximum group size imbalance: ratio of the larger group to the smaller group <4:1 (88). A study will be excluded if any of the following criteria are present: 1) Number of participants in the smallest group <5. 2) CV <0.1 or >0.95, and 3) Ratio of the larger to the smallest group >20:1. These criteria were derived from the principles outlined in the Cochrane Handbook, relevant methodology papers, and analogous statistical literature to ensure the robustness of the tertile-based calculations (66, 81, 87, 89).

If a study provides both adjusted and unadjusted effect sizes for a risk factor, we will only use the adjusted effect size in analysis. Additionally, we will report the mean differences between groups with and without RS for each continuous prognostic factor.

### Studies with Overlapping Populations

All included studies will be described in the patient characteristics table and their data will be used for descriptive analyses. However, to avoid potential bias and inflation of the statistical significance due to data duplication, only one study from each set of studies with overlapping populations will be included in the meta-analysis. Overlapping populations will be identified based on a careful examination of author information (including affiliations), the time period of patient recruitment, and, if available, details of the study setting (e.g., specific hospitals or clinics). When overlapping populations are identified, we will select the study with the largest sample size, assuming the smaller sample is a subset of the larger study (81, 90, 91).

### Biochemical Risk Factors

Serum total calcium and serum ionized calcium will be included in the same unadjusted odds ratio meta-analysis using the tertile approach. If a study reports both ionized and total calcium, only ionized calcium will be used, as it is considered a more accurate measure (92). Similarly, the uOR for 25-hydroxy vitamin D and 1-25 dihydroxy vitamin D will be meta-analyzed together and 25-hydroxy vitamin D will be prioritized over 1-25 dihydroxy vitamin D if both are available in a study, as 25-hydroxy vitamin D represents a more accurate measure of body’s vitamin D stores (93–95).

For the analysis of mean differences in biochemical factors between patients with and without RS, serum total calcium and serum ionized calcium will be analyzed separately. However, a single certainty of evidence recommendation will be provided for both, as they represent closely related clinical concepts and because this analysis is exploratory (96). The same analytic approach will be employed for mean differences and certainty of evidence reporting for 25-hydroxy vitamin D and 1-25 dihydroxy vitamin D.

## 11. Subgroup Analyses and Meta-regression

### Subgroup Analyses

To assess heterogeneity, we will conduct the following subgroup analyses:

1. **By Presence of Osteoporosis:** Studies with ≥20% will be compared with those with <20% of participants having osteoporosis (cutoff chosen based on global prevalence)(97). We expect a stronger association between risk factors and RS in patients with osteoporosis (98).
2. **By Study Design:** Cohort and case-control will be compared with cross-sectional studies. We expect a stronger association in cross-sectional studies as they tend to have higher bias (99, 100).
3. **By treatment with thiazide diuretics**: within patients treated with thiazide diuretics, we expect weaker association between the risk factors and RS as thiazide diuretics lower urinary calcium and potentially RS risk (101).

### Meta-regression

1. **By mean serum calcium in each study**: We hypothesize that the magnitude of the association between the risk factors and RS will be weaker in studies with higher mean serum calcium levels. This is because, at higher average serum calcium levels within a study population, hypercalcemia is likely to be a more dominant factor in stone formation, potentially overshadowing the influence of other risk factors(102, 103).
2. **By proportion of females in each study:** We hypothesize that the magnitude of the association between other risk factors and RS will be weaker in studies with a smaller difference in the proportion of females between the RS and no-RS groups. This indirect assessment is based on the assumption that a smaller difference in the proportion of females between the groups might reflect a population where sex-related factors play a less prominent role in modifying the influence of other risk factors on renal stone formation (104).

### Evaluating the Credibility of Subgroup & Meta-regression Analyses

Two reviewers will independently grade the credibility of each subgroup effect in duplicate using the Instrument for the Credibility of Effect Modification Analyses (ICEMAN) criteria (105). Disagreements will be resolved through discussion between the reviewers and the third reviewer will adjudicate unresolved disagreements. Credibility ratings will be provided assigned as follows: 1-Very Low: all responses are “definitely no” or “probably no. 2-Low: at least two responses are “definitely no.” 3-Moderate: one “definitely no” or two “probably no” responses. 4-High: no responses are “definitely no” or “probably no” (106).

## 12. Sensitivity Analyses

To confirm the robustness of our findings, we will conduct the following sensitivity analyses: 1) To assess the effect of **consistent covariate selection**, we will only include studies that adjust for at least serum calcium and either age or sex. 2) To examine the influence of **study design**, we will restrict the analysis to cohort and case-control studies, as these designs allow for stronger inferences about associations compared to cross-sectional studies (107). 3) To evaluate the impact of **risk of bias**, we will restrict to studies with low or moderate risk of bias. If a clinically significant difference is found between the primary or secondary analysis and their corresponding sensitivity analyses, we will present the primary analysis and rate down the certainty of evidence.

## 13. Reporting Bias

For analyses including ≥10 studies, we will conduct a primary assessment using funnel plots, the trim-and-fill method, Egger’s test for continuous variables, and the Harbord test for dichotomous variables (108–110). To complement these assessments, a secondary assessment will be performed using the Doi plot and Luis Furuya-Kanamori (LFK) index, with values within ±1 interpreted as indicative of no asymmetry (111, 112). In instances where the primary and secondary assessments yielded discrepant results, Begg’s test will be employed to provide additional clarification (113, 114). The final determination of publication bias will be based on the consensus of multiple analyses, prioritizing methods with greater sensitivity and reliability for the specific context (110, 115–117). For analyses with < 10 studies, formal assessments of publication bias will not be conducted due to limited statistical power. Instead, publication bias will be considered “unlikely” if small-study effects are improbable, based on the comprehensiveness of the search strategy and expert knowledge (109, 110, 118).

## 14. Confidence in Cumulative Evidence

Two independent reviewers will assess the overall quality of evidence for each risk factor’s association with the outcomes using the Grading of Recommendations Assessment, Development, and Evaluation (GRADE) framework (119). The assessment will be adapted to align with GRADE recommendations for prognostic factor studies (120).

## 15. Ethics Approval and Consent to Participate

We plan to conduct a systematic review and meta-analysis which will use data from previously published studies and will not collect new data from participants. Therefore, ethics approval and consent to participate will not be required.

## DISSEMINATION OF INFORMATION

Findings will be disseminated through presentations at scientific meetings and published through at least one peer-reviewed manuscript.

## DISCUSSION

To our knowledge, this is the first systematic review and meta-analysis designed to synthesize and quantify the existing evidence on key risk factors for RS in the setting of PHPT. By addressing the unique demographic and metabolic factors specific to PHPT, this study aims to resolve inconsistencies in the literature and identify PHPT-specific risk factors that may differ from those in the general population. Identifying these risk factors will enable clinicians to more effectively risk-stratify patients with PHPT, facilitating targeted preventive measures, closer monitoring, and timely parathyroidectomy for high-risk individuals. Ultimately, this could reduce the burden of RS, including emergency department visits, renal failure, and associated healthcare costs (4–7, 11, 12).

### Limitations

We anticipate that many relevant studies will use a cross-sectional design. While combining cross-sectional, cohort, and case-control studies in the meta-analysis will enhance precision by increasing the number of included studies (121), the lack of a temporal component in cross-sectional studies will limit our ability to determine whether these risk factors serve as predictors. To address this limitation, we will conduct sensitivity analyses restricted to cohort and case-control studies. Additionally, pooling adjusted estimates that include different covariates will augment the overall sample size but reduce direct comparability of summary estimates between studies and complicate interpretation of effect size magnitude (54, 122, 123). To address these limitations, we will conduct sensitivity analyses restricted to studies adjusting for key confounders.

## Data Availability

No datasets were generated or analysed during the current study. All relevant data from this study will be made available upon study completion.

## AUTHOR CONTRIBUTION

Conceptualization: Mohammad Jay

Methodology: Mohammad Jay, Antoine Eskander, Lorraine Lipscombe, Rui Fu

Project administration: Mohammad Jay, Sorina Andrei

Supervision: Antoine Eskander, Lorraine Lipscombe

Validation: Mohammad Jay, Lorraine Lipscombe, Antoine Eskander, Sorina Andrei, Peter

Hoang, Roland Jones, Hussein Samhat

Writing – original draft: Mohammad Jay

Writing – review & editing: Mohammad Jay, Sorina Andrei, Peter Hoang, Hussein Samhat, Roland Jones, Rui Fu, Lorraine Lipscombe, Antoine Eskander

## ACKNOWLEDGEMENT

We extend our gratitude to Dr. Neill Adhikari and Dr. Danny Weisz for their contributions to the methodological development of this project. We also sincerely thank Ms. Genevieve Gore for her support in developing the draft search strategy.

## SUPPORTING INFORMATION

**<u>S1. Appendix:</u> Preferred Reporting Items for Systematic review and Meta-Analysis Protocols 2015 checklist**

**<u>S2. Appendix</u>: Draft search strategy**

**<u>S3. Appendix:</u> Variables for Data Extraction**

