## Supporting Document for "Risk factors for renal stone development in adults with primary hyperparathyroidism: A protocol for a systematic review and meta-analysis"

**Heading: SUPPORTING INFORMATION**

**Sub-heading: S1. Appendix**

**Preferred Reporting Items for Systematic review and Meta-Analysis Protocols 2015 checklist: recommended items to address in a systematic review protocol***

| Section and topic | Item No | Checklist item | Page |
| --- | --- | --- | --- |
| ADMINISTRATIVE INFORMATION | | |  |
| Title: |  |  | 1 |
| Identification | 1a | Identify the report as a protocol of a systematic review | 1 |
| Update | 1b | If the protocol is for an update of a previous systematic review, identify as such | NA |
| Registration | 2 | If registered, provide the name of the registry (such as PROSPERO) and registration number | NA |
| Authors: |  |  | 1 |
| Contact | 3a | Provide name, institutional affiliation, e-mail address of all protocol authors; provide physical mailing address of corresponding author | 1 |
| Contributions | 3b | Describe contributions of protocol authors and identify the guarantor of the review | 1 |
| Amendments | 4 | If the protocol represents an amendment of a previously completed or published protocol, identify as such and list changes; otherwise, state plan for documenting important protocol amendments | N/A |
| Support: |  |  | NA |
| Sources | 5a | Indicate sources of financial or other support for the review | NA |
| Sponsor | 5b | Provide name for the review funder and/or sponsor | NA |
| Role of sponsor or funder | 5c | Describe roles of funder(s), sponsor(s), and/or institution(s), if any, in developing the protocol | NA |
| INTRODUCTION | | |  |
| Rationale | 6 | Describe the rationale for the review in the context of what is already known | 4-5 |
| Objectives | 7 | Provide an explicit statement of the question(s) the review will address with reference to participants, interventions, comparators, and outcomes (PICO) | 5-6 |
| METHODS | | |  |
| Eligibility criteria | 8 | Specify the study characteristics (such as PICO, study design, setting, time frame) and report characteristics (such as years considered, language, publication status) to be used as criteria for eligibility for the review | 6-8 |
| Information sources | 9 | Describe all intended information sources (such as electronic databases, contact with study authors, trial registers or other grey literature sources) with planned dates of coverage | 8-9 |
| Search strategy | 10 | Present draft of search strategy to be used for at least one electronic database, including planned limits, such that it could be repeated | 9 & Appendix II |
| Study records: |  |  |  |
| Data management | 11a | Describe the mechanism(s) that will be used to manage records and data throughout the review | 9-11 |
| Selection process | 11b | State the process that will be used for selecting studies (such as two independent reviewers) through each phase of the review (that is, screening, eligibility and inclusion in meta-analysis) | 9 |
| Data collection process | 11c | Describe planned method of extracting data from reports (such as piloting forms, done independently, in duplicate), any processes for obtaining and confirming data from investigators | 10 |
| Data items | 12 | List and define all variables for which data will be sought (such as PICO items, funding sources), any pre-planned data assumptions and simplifications | 10-11 |
| Outcomes and prioritization | 13 | List and define all outcomes for which data will be sought, including prioritization of main and additional outcomes, with rationale | 8 |
| Risk of bias in individual studies | 14 | Describe anticipated methods for assessing risk of bias of individual studies, including whether this will be done at the outcome or study level, or both; state how this information will be used in data synthesis | 11 |
| Data synthesis | 15a | Describe criteria under which study data will be quantitatively synthesised | 12-15 |
|  | 15b | If data are appropriate for quantitative synthesis, describe planned summary measures, methods of handling data and methods of combining data from studies, including any planned exploration of consistency (such as I^2^, Kendall’s τ) | 12-15 |
|  | 15c | Describe any proposed additional analyses (such as sensitivity or subgroup analyses, meta-regression) | 15-17 |
|  | 15d | If quantitative synthesis is not appropriate, describe the type of summary planned | 14-15 |
| Meta-bias(es) | 16 | Specify any planned assessment of meta-bias(es) (such as publication bias across studies, selective reporting within studies) | 17-18 |
| Confidence in cumulative evidence | 17 | Describe how the strength of the body of evidence will be assessed (such as GRADE) | 18 |

*From: Shamseer L, Moher D, Clarke M, Ghersi D, Liberati A, Petticrew M, Shekelle P, Stewart L, PRISMA-P Group. Preferred reporting items for systematic review and meta-analysis protocols (PRISMA-P) 2015: elaboration and explanation. BMJ. 2015 Jan 2;349(jan02 1):g7647.*

**Subheading: S2. Appendix**

**Draft search strategy**

| **Ovid Medline(R) All <1946 to September 26, 2024>** ^1^ | | |
| --- | --- | --- |
| **ID** | **Search** | **Hits** |
| 1 | Hyperparathyroidism, Primary/ | 4064 |
| 2 | Parathyroid Glands/ | 13342 |
| 3 | exp Parathyroid Hormone/ | 32084 |
| 4 | Parathyroid Neoplasms/ | 8628 |
| 5 | exp Multiple Endocrine Neoplasia/ | 5770 |
| 6 | ((primary adj1 hyperparathyroid*) or parathyroid hormone* or parathyroid gland* or parathyroid neoplasm* or (MEN adj ("1" or "2" or I or II)) or MEN1 or MENI or MEN2 or MENII or MEN syndrome* or multiple endocrine neoplas*).tw,kf. | 59673 |
| 7 | or/1-6 | 78202 |
| 8 | exp Nephrolithiasis/ | 23794 |
| 9 | exp Urolithiasis/ | 43805 |
| 10 | Nephrocalcinosis/ | 2281 |
| 11 | Nephrosclerosis/ | 1789 |
| 12 | (((kidney? or renal or staghorn or ureter* or urinary or vesical) adj2 (calcification* or calcul* or concret* or gravel or lithiasis or stone*)) or cystolith* or nephrolith* or nephrocalcinos* or nephrosclero* or urolith* or ureterolith* or vesicolith*).tw,kf. | 53226 |
| 13 | or/8-12 | 66502 |
| 14 | 7 and 13 | 2469 |
| **Central (Cochrane Library/Wiley) <inception to Sept 26, 2024>** | | |
| 1 | [mh ^"Hyperparathyroidism, Primary"] OR [mh ^"Parathyroid Glands"] OR [mh "Parathyroid Hormone"] OR [mh ^"Parathyroid Neoplasms"] OR [mh "Multiple Endocrine Neoplasia"] OR ((primary NEAR/1 hyperparathyroid*) OR ("parathyroid" NEAR/2 hormone*) OR ("parathyroid" NEAR/2 gland*) OR ("parathyroid" NEAR/2 neoplasm*) OR (MEN NEXT (1 OR 2 OR I OR II)) OR MEN1 OR MENI OR MEN2 OR MENII OR ("MEN" NEAR/2 syndrome*) OR ("multiple endocrine" NEAR/2 neoplas*)) | 5937 |
| 2 | ([mh Nephrolithiasis]) OR ([mh Urolithiasis]) OR ([mh ^Nephrocalcinosis]) OR ([mh ^Nephrosclerosis]) OR ((((kidney? OR renal OR staghorn OR ureter* OR urinary OR vesical) NEAR/2 (calcification* OR calcul* OR concret* OR gravel OR lithiasis OR stone*)) OR cystolith* OR nephrolith* OR nephrocalcinos* OR nephrosclero* OR urolith* OR ureterolith* OR vesicolith*) | 6703 |
|  | ,ab,kw) |  |
| 3 | #1 AND #2 in Trials | 116 |
| **Embase Classic+ Embase <1947 to 2024 September 26>** | | |
| 1 | exp *primary hyperparathyroidism/ | 8364 |
| 2 | exp *parathyroid gland/ | 7797 |
| 3 | *parathyroid hormone/ | 20601 |
| 4 | exp *multiple endocrine neoplasia/ | 4012 |
| 5 | ((primary adj1 hyperparathyroid*) or parathyroid hormone* or parathyroid gland* or parathyroid neoplasm* or (MEN adj ("1" or "2" or I or II)) or MEN1 or MENI or MEN2 or MENII or MEN syndrome* or multiple endocrine neoplas*).tw,kf. | 82090 |
| 6 | or/1-5 | 90503 |
| 7 | exp urolithiasis/ | 86220 |
| 8 | kidney calcification/ | 7738 |
| 9 | exp nephrosclerosis/ | 5018 |
| 10 | (((kidney? or renal or staghorn or ureter* or urinary or vesical) adj2 (calcification* or calcul* or concret* or gravel or lithiasis or stone*)) or cystolith* or nephrolith* or nephrocalcinos* or nephrosclero* or urolith* or ureterolith* or vesicolith*).tw,kf. | 83231 |
| 11 | or/7-10 | 114088 |
| 12 | 6 and 11 | 3340 |
| 13 | limit 12 to conference abstract status | 964 |
| 14 | \| 12 not 13 | 3349 |

1- Two main concepts were included in the search strategy: primary hyperparathyroidism, and renal stone.

**Subheading: S3. Appendix**

**S3. Variables for Data Extraction**

**Table 1. Basic Study Details**

| Study ID | First Author's Last Name | Publication Year | Study Design | Country of Origin | Sample Size | Setting | Data Source | Follow-up Duration | Loss to Follow-up | Funding Source |
| --- | --- | --- | --- | --- | --- | --- | --- | --- | --- | --- |

**Table 2. Patient Demographics and Clinical History**

| Age | Sex | Time of PHPT Diagnosis in Relation to RS Development | Criteria for PHPT Diagnosis | Presence of RS (Specify if RS is Incident or Recurrent) | Criteria for RS Diagnosis |
| --- | --- | --- | --- | --- | --- |

**Table 3. Biochemical Risk Factors**

| Serum Total Calcium | Serum Corrected Calcium | Serum Ionized Calcium | Serum PTH | 24-hour Urine Calcium | Units of Measurement for Biochemical Variables | Time Recorded in Relation to RS Development |
| --- | --- | --- | --- | --- | --- | --- |

**Table 4. Potential Confounders**

| Osteoporosis | Serum Creatinine | Serum eGFR | 24-hour Urine Completion | Treatment with Thiazide Diuretics |
| --- | --- | --- | --- | --- |

**Table 5. Effect Sizes and Descriptive Statistics**

| Adjusted Effect Size (OR, RR, HR) | Unadjusted Effect Size (OR, RR, HR) | Mean Difference | Measures of Variance | Descriptive Statistics (mean, median, frequency) | P-value | Variables Used for Adjustment |
| --- | --- | --- | --- | --- | --- | --- |
